# Opioid misuse is mediated by affective states in chronic pain patients

**DOI:** 10.1101/2025.04.14.25325813

**Authors:** Jesús D. Lorente, Francisco Molins, Félix Padilla, Diana S. Herrera, Vicente Monsalve Dolz, Ana Mínguez, Jose De Andres, Miguel Ángel Serrano, Lucía Hipólito

## Abstract

Chronic pain is closely associated with psychological comorbidities, including anxiety and depression, as well as substance use disorders, particularly alcohol use disorder and opioid use disorder. Recent research highlights the bidirectional nature of these relationships, suggesting that each of these variables can contribute to the onset and exacerbation of the others. The present study aims to examine whether negative affective states associated with chronic pain are linked to risk-related patterns of opioid and/or alcohol consumption in individuals suffering from chronic pain. To this end, a comprehensive battery of standardized instruments was administered to assess subjective pain intensity, anxious-depressive symptomatology, and substance use risk patterns for opioids and alcohol. The findings reveal a strong positive correlation between subjective pain levels, anxious-depressive symptoms, and opioid risk-taking behaviors. Notably, individuals with elevated scores on measures of anxiety and depression reported higher levels of subjective pain and demonstrated greater risk-taking behavior related to opioid use, compared to those with lower symptomatology scores. In conclusion, this study underscores chronic pain as a significant risk factor for the development of anxious-depressive disorders and opioid misuse. These findings highlight the importance of integrating psychological and substance use assessments in the management of chronic pain to inform targeted interventions and preventative strategies.

## Introduction

Chronic pain is a complex and burdensome condition that often coexists with psychological comorbidities, particularly anxiety, depression, and anhedonia [2,21,37]. Patients living with chronic pain are more than twice as likely to experience mood and anxiety disorders compared to those without pain [2,46,61]. This strong association underscores the need to consider chronic pain not just as a sensory experience but also as a risk factor for developing significant affective disturbances.

Evidence from both clinical populations and animal models highlights the profound impact of chronic pain on the brain’s mesocorticolimbic system, an area critically involved in emotion regulation and reward processing [3,6,40,44,47,48]. Pain-induced neuroadaptations in this system are associated with the emergence of negative affective states, which may contribute to maladaptive coping behaviors, such compulsive alcohol drinking or opioid-analgesic medication misuse.

In primary care, approximately 51% of patients with chronic pain report using drugs, whether prescribed, illicit, or misused, to manage their symptoms [1]. In a recent study, among patients with opioid use disorder (OUD), over 60% experienced chronic pain prior to developing OUD [27]. In fact, long-term opioid use is related to problems such as addiction and overdose and deterioration of physical and mental health problems [2,17]. Pain-induced emotional distress appears to play a central role in the initiation and relapse of substance use, as individuals may turn to opioids or alcohol to self-medicate their psychological suffering [60,63]. Indeed, different studies have related the presence of depression and anxiety to higher rates of OUD in chronic pain patients [4,7,53], as well as this effect of pain-induced negative affective states have been demonstrated for alcohol misuse and relapse [14,29,64].

Importantly, sex differences have emerged as a critical factor in pain perception, emotional response, and vulnerability to addiction [10,11,19,23,25,43,45,50–52,55,57]. Some research has shown that women may be more responsive to the adverse emotional effects of pain, which can affect their physical motor function [25]. These differences must be considered to tailor effective interventions.

Given the pressing opioid crisis and the widespread burden of chronic pain, it is crucial to better understand how emotional distress contributes to substance misuse in this population with the final objective of rational use of pain medications minimizing risks. In this study, we investigate the mediating role of negative affect—specifically anxiety, depression, and anhedonia—in the relationship between chronic pain and opioid or alcohol misuse. We further explore potential sex differences in these pathways. Our goal is to provide clinical strategies for early identification of patients at risk, enabling targeted, interdisciplinary interventions that reduce the reliance on opioids and improve overall patient care.

## Material and methods

### Participants

101 participants were recruited from the multidisciplinary treatment of pain unit of the *Consorcio Hospital General Universitario de Valencia,* although 4 participants did not complete all the measures taken in this protocol and were excluded from the study. Thus, our sample was finally comprised by a total of 97 participants (age: *M* = 57.77, *SD* = 13.56; women: 69.1%). All of them were diagnosed with some chronic pain pathology, such as neuropathic pain, fibromyalgia, arthritis, osteoarthritis or others. In addition, they met the following inclusion criteria: be a chronic pain patient (no migrane and cancer-pain) for at least 6 months, preserved the cognitive ability to complete the different scales and have opioids prescribed for the treatment of pain.

### Procedure

All participants read and signed informed consent and completed the battery of questionnaires, which included socio-economic questions, such as age, gender, or educational level. This study was approved by the Ethics Research Committee of the University of Valencia and the Clinical Research Ethics Committee of the Consorcio Hospital General Universitario de Valencia in accordance with the ethical standards of the 1969 Declaration of Helsinki.

### Alcohol Use Disorders Identification Test (AUDIT)

The Alcohol Use Disorders Identification Test [5] is a screening tool developed by the World Health Organization to screen individuals for risky or hazardous alcohol consumption or alcohol use disorder. It consists of 10 items covering three domains: alcohol consumption, drinking behavior/dependence, and adverse consequences of drinking. Scores range from 0 to 40, with a threshold score of 8 generally indicating a potential alcohol problem.

### Current Opioid Misuse Measure (COMM)

The Current Opioid Misuse Measure [13] is a self-report assessment tool designed to identify risk for aberrant medication-related behaviors among chronic pain patients prescribed opioids. It comprises 17 items, rated on a scale from 0 (“never”) to 4 (“very often”), reflecting behaviors over the past 30 days, with higher scores indicating a greater risk of opioid misuse. A score of 9 or higher is generally considered indicative of pathological consumption.

### Beck Anxiety Inventory (BAI)

The Beck Anxiety Inventory [8] is a self-report inventory for measuring the severity of anxiety. The scale consists of 21 items, each describing a common symptom of anxiety. The respondent is asked to rate the extent to which each symptom has affected them over the past week on a 4-point scale, from 0 (symptom absent) to 3 (severe symptoms). Scoring is achieved by adding the highest ratings for all 21 items and the higher the scores, the greater symptom severity. In addition to the total score, participants were divided into the following categories: minimal anxiety (0–7), mild anxiety (8–15), moderate anxiety (16–25), and severe anxiety (26–63).

### Beck Depression Inventory (BDI) II fast screen

The Beck Depression Inventory [9] is a 7-item self-reporting questionnaire for evaluating the severity of depression in normal and psychiatric populations. Each item is rated on a 4-point scale from 0 (symptom absent) to 3 (severe symptoms). Affective, cognitive, somatic, and vegetative symptoms are covered, reflecting the DSM-IV criteria for major depression. Scoring is achieved by adding the highest ratings for all 7 items. Higher scores indicate greater symptom severity. In non-clinical populations, scores above 3 indicate depression. Furthermore, participants were divided into the following categories: minimal depression (0–3), mild depression (4–8), moderate depression (9–12), and severe depression (13–21).

### Snaith-Hamilton Pleasure Scale (SHAPS)

The Snaith-Hamilton Pleasure Scale [59] is a self-report questionnaire designed to assess anhedonia, the diminished ability to experience pleasure, which is a core symptom of depression. The SHAPS consists of 14 items, each describing a pleasurable experience (e.g., “I would enjoy being with family or close friends”). Respondents are asked to rate how much they agree with each statement over the past few days on a 4-point scale: “Strongly Disagree,” “Disagree,” “Agree,” and “Strongly Agree.” Each item is scored dichotomously as 0 (Agree/Strongly Agree) or 1 (Disagree/Strongly Disagree), with total scores ranging from 0 to 14. A score of 3 or higher is generally considered indicative of significant anhedonia, so this cut-off was used to differentiate between those with and without severe anhedonia.

### Visual Analogue Scale (VAS) of Pain

Subjective pain ratings were made using a Visual Analogue Scale (VAS) [18]including anchors of “no pain sensation” and “most intense pain imaginable.” The VAS was instantiated on paper as a 10 cm horizontal line with the two anchors, and participants indicated their pain level by marking a spot on the line. To obtain a numerical score (0–10) for this rating, a ruler was used to measure the distance (in mm). This scale was used to measure pain experienced over the last month (last-month VAS) and on the day of the assessment (day-VAS). 3 to 7 are considered moderate pain scores, while 8 to 10 would indicate severe pain.

### Statistical analyses

Outliers were identified using the 2.5 standard deviations method and normality was assessed using Kolmogorov-Smirnoff with Lilliefors correction. Exploratory analyses were conducted to study the distribution of variables and their interrelations by sex. Subsequently, MANOVAs were performed to compare subjective pain (last month-VAS and day-VAS) and opioid consumption (COMM) across categories of anxiety (BAI), depression (BDI), and anhedonia (SHAPS). Complementary clustering analyses were conducted to examine how participants were grouped based on their levels of anxiety, depression, and anhedonia. These clusters were then analyzed to identify differences in pain and opioid use, as well as to explore the relationships between these variables within the clusters. Finally, mediation analyses were performed to investigate whether anxiety, depression, and anhedonia mediate the association between subjective pain and opioid consumption. The Sobel test was utilized to check the statistically significance of the mediation models.

The significance level (α) was set at .05, and the partial eta square (η^2^_p_) indicated the effect size. All analyses were conducted using IBM SPSS Statistics 25.

## Results

### Exploratory analyses

Table 1 shows the means and standard deviations of all the variables included in this work. As observed, our sample possessed a moderate educational level (3.35 out of 6) and an average duration of 10.79 years experiencing their pain-related condition. Furthermore, low scores were obtained in the AUDIT, indicating a negligible alcohol consumption. On the other hand, the mean COMM scores revealed problematic opioid consumption in the sample (>9). Anxiety levels were moderate (between 16 and 25), while depression levels were minimal (between 0 and 13), and anhedonia did not exceed the cut-off score of 3 that would indicate a problematic level. Finally, the VAS scores (last month and day) indicated a moderate level of subjective pain, bordering on intense.

**Table 1.** Descriptive statistics.

|  | N = 97 |
| --- | --- |
| Age | 57.77 ± 13.55 |
| Educational level | 3.35 ± 1.26 |
| Chronic pain duration (years) | 10.79 ± 10.88 |
| AUDIT | 1.70 ± 2.15 |
| COMM | 13.16 ± 10.71 |
| BAI | 18.79 ± 13.69 |
| BDI | 5.47 ± 4.78 |
| SHAPS | 1.33 ± 2.10 |
| Last-month VAS of pain | 7.79 ± 1.69 |
| Day VAS of pain | 6.95 ± 2.18 |
| Mean ± SD; educational level ranges from 1 (without studies) to 6 (University studies) | |

A correlation analysis was then performed for all the variables obtained with the different tests and questionnaires of the study (COMM, AUDIT, BDI, BAI, SHAPS, and last day and last month VAS). Figure 1A shows the correlation matrix focusing on the main seven variables on which the study is targeted.

**Figure 1.**
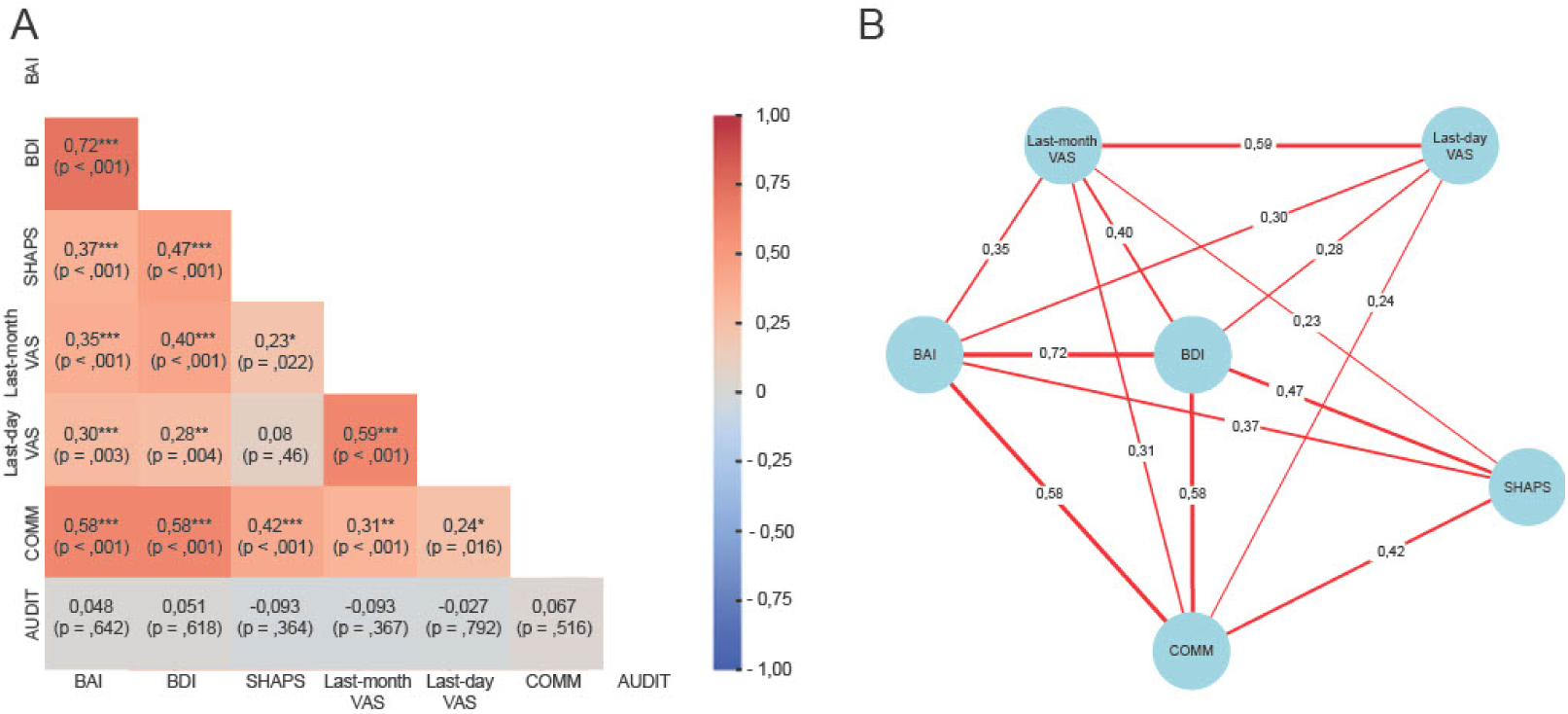
Heat map and Network of the correlations between BAI, BDI, SHAPS, VAS, and COMM. (A) Heat map featuring a color gradient based on the intensity of correlation. Values in each cell represent Pearson’s correlation coefficient, and asterisks denote significant correlations (*** significant at .001 level; ** significant at .01 level; * significant at .05 level). (B) Network that enhances the comprehension of the previous heat map by visually depicting the connections between variables. The edges vary in thickness in accordance with the strength of their correlation

As can be seen, almost all variables were significantly related to each other, apart from the variable related to risk patterns of alcohol consumption (AUDIT), that was not related to any of the other 6 variables (values of the Pearson’s correlation coefficient and p-values are detailed in the Heatmap of the Figure 1A). A strong correlation was found between anxious (BAI) and depressive (BDI) symptomatology, and medium correlations were observed between depressive or anxious symptomatology and anhedonia (SHAPS). Both BAI and BDI showed correlations ranging from low to medium with subjective pain assessed with the Last-month VAS and Day VAS, as well as strong, positive correlations with the opioid consumption (COMM). Additionally, a medium correlation was observed between anhedonia and opioid consumption. Finally, significant correlations were also observed between the Last-month VAS or Day VAS and COMM, with these relationships being of low to medium strength (Figure 1A and 1B).

These correlations were also separately tested for each gender.

As seen in Figure 2, women maintained similar relations between the main variables, also showing no correlation between AUDIT and the rest of the variables. Again, the strongest correlation was found between anxious and depressive symptomatology. Strong relationships were also observed between both anxious and depressive symptomatology with opioid consumption, and a medium correlation was observed between anhedonia and opioid consumption. Anxious and depressive symptomatology also showed correlations ranging from low to medium with subjective pain. Finally, subjective pain was significantly correlated to opioid consumption, with relationships being of low to medium strength.

**Figure 2.**
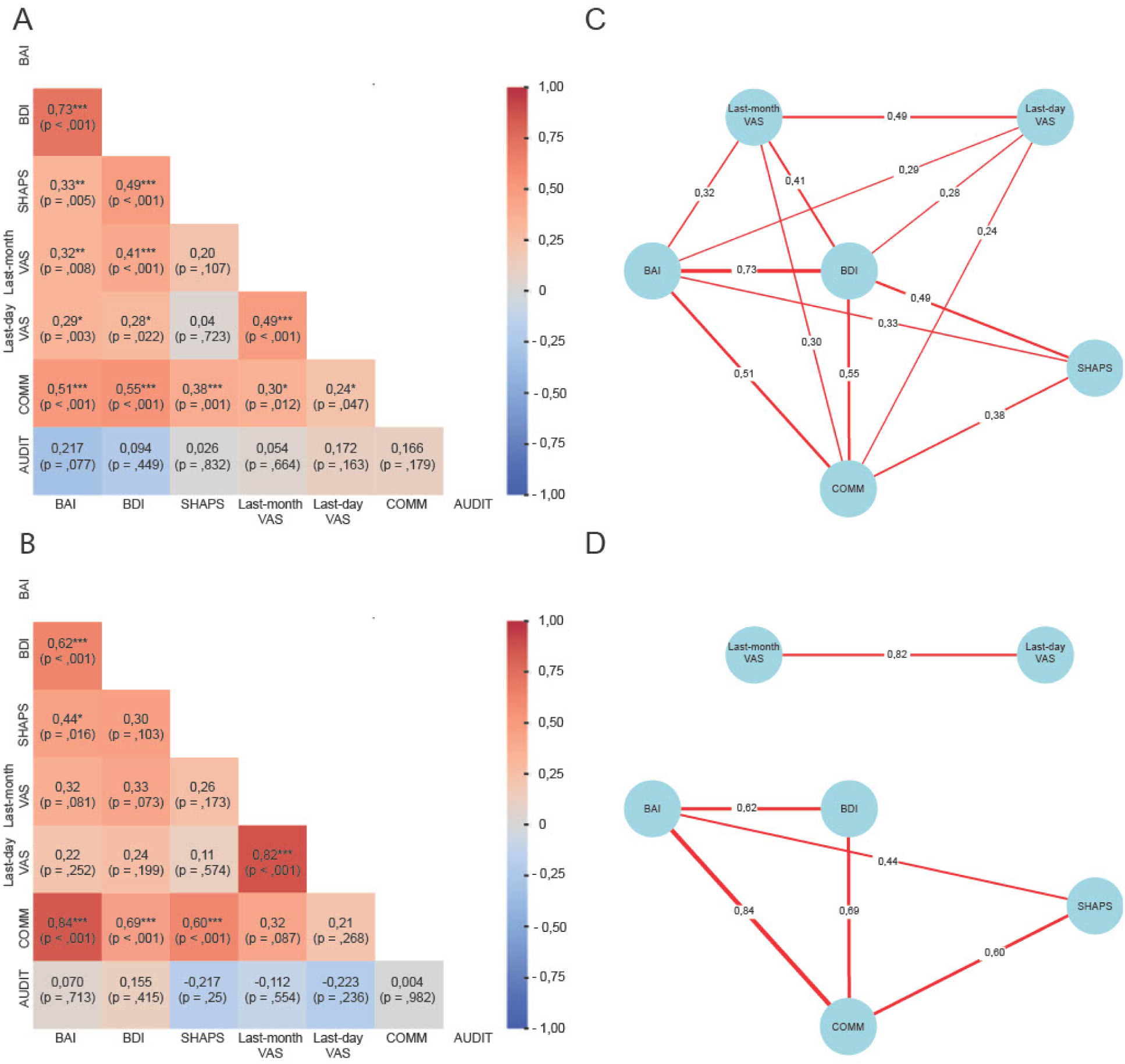
Heat map and Network of the correlations between BAI, BDI, SHAPS, VAS, and COMM, by gender. Heat map for women (A) and men (B) featuring a color gradient based on the intensity of correlation. Values in each cell represent Pearson’s correlation coefficient, and asterisks denote significant correlations (*** significant at .001 level; ** significant at .01 level; * significant at .05 level). Network that enhances the comprehension of the previous heat map by visually depicting the connections between variables. (C) for women; (D) for men. The edges vary in thickness in accordance with the strength of their correlation.

Regarding men, as in the case of women, the AUDIT variable did not correlate with any of the variables. In addition, some of the correlations that were found for the whole population and for women were not shown for men. The analysis detected strong relations between depressive and anxious symptomatology. Interestingly, both anxious and depressive symptomatology had strong correlations with opioid consumption. Anhedonia also showed a medium correlation with opioid consumption. However, the correlations between anxious or depressive symptomatology and subjective pain were not significant, although they exhibited a statistical trend (p values ranging from .074 to .087). In the same line, no correlations were found between subjective pain and opioid consumption.

### Differences in subjective pain and opioid consumption by intensity of anxiety and depression

Since the pattern of alcohol consumption variable is not significantly related to any variable, as it can be observed in the previous section, it has not been included in the following analyses.

First, we divided the participants according to the intensity of their anxiety assessed with BAI. By means of a MANOVA, and including the gender variable, we compared the levels of subjective pain and opioid consumption according to the groups created. Neither gender nor its interaction with anxiety significantly explained the variance of the variables analyzed. However, anxiety explains the variance of COMM between group and pain perception during last month, but not pain perception during the same day. As can be seen in Table 2, *post hoc* comparisons revealed that those participants with severe anxiety presented greater pain in the last month with respect to minimal and mild anxiety. Moderate anxiety presented pain levels that would be midway between the groups with lower anxiety and those with higher anxiety. On the other hand, although no significant differences in COMM score were observed between the moderate and severe anxiety groups, both groups significantly scored higher in the COMM than those with minimal or mild anxiety.

**Table 2.** Differences in subjective pain (Last month and day VAS), and opioid consumption (COMM) by anxiety intensity.

| | Minimal anxiety (19) | Mild anxiety (34) | Moderate anxiety (17) | Severe anxiety (27) | *F* | df intra | df error | *p*-value | $\eta_p^2$ |
| --- | --- | --- | --- | --- | --- | --- | --- | --- | --- |
| Last-month VAS | 7.28 ± 1.98<sup>a</sup> | 7.20 ± 1.94<sup>a</sup> | 8.14 ± 1.02<sup>ab</sup> | 8.68 ± 0.93<sup>b</sup> | 2.833 | 3 | 89 | .043* | .087 |
| Day VAS | 6.78 ± 2.12<sup>a</sup> | 6.38 ± 2.24<sup>a</sup> | 6.47 ± 1.65<sup>a</sup> | 8.11 ± 2.09<sup>a</sup> | 1.964 | 3 | 89 | .125 | .062 |
| COMM | 5.58 ± 5.39<sup>a</sup> | 9.15 ± 6.21<sup>a</sup> | 17.12 ± 11.13<sup>b</sup> | 21.07 ± 11.84<sup>b</sup> | 14.056 | 3 | 89 | <.001*** | .321 |

Mean ± *SD* by anxiety categories (n); df, degrees of freedom; groups with the same letter in superscript (<sup>a</sup> or <sup>b</sup>) do not differ significantly; different letters represent significant differences according to *post hoc* tests; \* significance at the .05 level; \*\* significance at the .01 level; \*\*\* significance at the .001 level

Moreover, this analysis was replicated by comparing the categories according to the intensity of depression (see Table 3). Again, neither gender nor its interaction with depression significantly predicted subjective pain or opioid use. However, depression explains the variance of COMM between group and pain perception during last month, but not pain perception during the same day. On the one hand, regarding subjective pain referred to last month, significant differences were observed between groups, however, post hoc tests did not detect any difference between the subjective level of pain depending on depressive symptomatology. On the other hand, subjective day pain, statistical tests showed no differences between any of the groups. Finally, in relation to COMM, the mild, moderate and severe depression groups showed higher scores on the COMM than the minimal depression group. In addition, higher scores were also observed for the moderate and severe depression groups compared to the mild depression group, with only the comparison between the mild and moderate groups being significant. Thus, it was possible to confirm that the intensity of depression influences subjective pain and the risk pattern of opioid use, although the sex of the participants and the interaction between sex and depression were not shown to be determining factors in these results.

**Table 3.** Differences in subjective pain (Last month and day VAS), and opioid consumption (COMM) by depression intensity.

| | Minimal<br>depression (41) | Mild<br>depression (33) | Moderate<br>depression (15) | Severe<br>depression (8) | *F* | df<br>intra | df<br>error | *p*-value | $\eta_p^2$ |
| --- | --- | --- | --- | --- | --- | --- | --- | --- | --- |
| Last-month VAS | 7,11 ± 1,91<sup>a</sup> | 7,98 ± 1,49<sup>a</sup> | 8,67 ± 0,83<sup>a</sup> | 8,94 ± 0,86<sup>a</sup> | 3,926 | 3 | 89 | ,024* | ,100 |
| Day VAS | 6,41 ± 2,19<sup>a</sup> | 6,99 ± 2,2<sup>a</sup> | 7,9 ± 2,04<sup>a</sup> | 7,88 ± 1,64<sup>a</sup> | 1,338 | 3 | 89 | ,267 | ,043 |
| COMM | 7,07 ± 5,30<sup>a</sup> | 13,00 ± 8,40<sup>b</sup> | 26,67 ± 11,41<sup>c</sup> | 19,75 ± 13,18<sup>bc</sup> | 16,593 | 3 | 89 | <,001*** | ,359 |

Mean ± *SD* by depression categories (n); df, degrees of freedom; groups with the same letter in superscript (<sup>a</sup> or <sup>b</sup>) do not differ significantly; different letters represent significant differences according to *post hoc* tests; \* significance at the .05 level; \*\* significance at the .01 level; \*\*\* significance at the .001 level

The last step was to compare the subjective pain and opioid consumption between non-severe and severe anhedonia (table 4). Gender and its interaction with anhedonia did not explain significant variance. However, anhedonia explains the variance of COMM between groups but not pain perception neither during the same day or during the last month. Both groups of anhedonia did not differ in their last month nor in their last day subjective pain. However, severe anhedonia showed a significant higher opioid misuse (COMM) score than the non-severe anhedonia.

**Table 4.** Differences in subjective pain (Last month and day VAS), and opioid consumption (COMM) by the presence of anhedonia.

| | Non-severe anhedonia | Severe anhedonia | *F* | df<br>intra | df<br>error | *p*-value | $\eta_p^2$ |
| --- | --- | --- | --- | --- | --- | --- | --- |
| Last-month VAS | 7.63 ± 1.79<sup>a</sup> | 8.59 ± 2.49<sup>a</sup> | 3,745 | 3 | 93 | ,06 | ,039 |
| Day VAS | 6,86 ± 2,12<sup>a</sup> | 7.35 ± 2,2<sup>a</sup> | ,231 | 3 | 93 | ,63 | ,002 |
| COMM | 11.61 ± 9.18<sup>a</sup> | 20.47 ± 14.29<sup>b</sup> | 7,934 | 3 | 93 | ,006** | ,079 |

Mean ± *SD* by depression categories (n); df, degrees of freedom; groups with the same letter in superscript (<sup>a</sup> or <sup>b</sup>) do not differ significantly; different letters represent significant differences according to *post hoc* tests; \* significance at the .05 level; \*\* significance at the .01 level; \*\*\* significance at the .001 level

### Anxiety, depression and anhedonia mediates between the relationship between subjective pain and risk of opioid misuse?

Given that subjective pain and COMM scores differed considering affective states, we further investigated whether the relationship between these variables was also modulated by this factor. Specifically, we conducted mediation analyses to examine whether anxiety, depression, and anhedonia mediated the relationship between pain and the scores obtained in the COMM (see Figure 3 to consult the requirements to consider that a variable mediates between two others). Since previous analyses highlighted that affective states had a greater effect on pain measured during the last month (rather than the last day), mediation analyses were conducted using last-month pain (Last-month VAS) as the independent variable. Sobel’s test was also conducted to test the significance of the mediation models.

**Figure 3.**
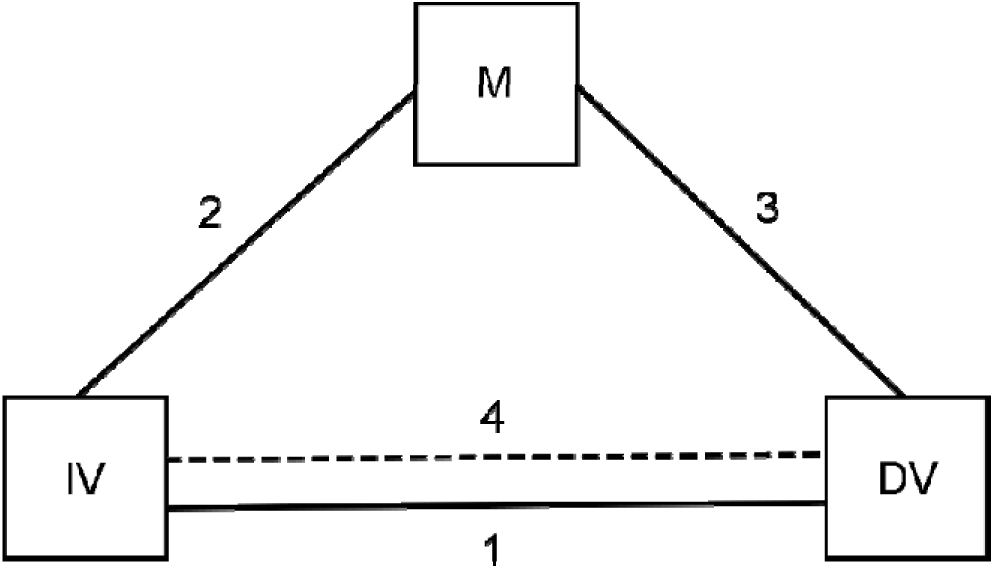
Mediation analysis flow (1) The independent variable (IV; subjective pain) and the dependent variable (DV; opioid consumption) must be linearly and significantly related; (2) the IV must be significantly related to the mediating variable (M; symptoms of anxiety or depression or anhedonia); (3) M must be significantly related to the DV; and (4) the relationship between the IV and the DV must weaken or disappear when the M is included in the model.

#### Anxiety symptoms mediate between subjective pain and opioid consumption

First, subjective pain was significantly associated with the COMM score, *B* = .06, *SE* = .23, *t*(95) = 2.46, *p* = .015. Second, subjective pain was also associated with the anxiety symptoms, *B* = 2.78, *SE* = .78, *t*(95) = 3.55, *p* = .001. Third, these symptoms were significantly associated with the COMM score, *B* = .46, *SE* = .06, *t*(95) = 7.11, *p* < .001. Finally, the relationship between the subjective pain and the COMM score disappeared (*B* = .70, *SE* = .56, *t*(94) = 1.24, *p* = .218), when controlling for anxiety, *B* = .44, *SE* = .07, *t*(94) = 6.28, *p* < .001. Furthermore, the Sobel test indicated that this mediation was significant, *z* = 3.1, *p* = .001.

This mediation model was replicated separately for women and men. In women, similar results were obtained, indicating that anxiety mediated the relationship between pain and COMM score (Sobel Test: *z* = 2.3, *p* = .021). Thus, subjective pain was significantly associated with the opioid consumption, *B* = 1.99, *SE* = .86, *t*(65) = 2.31, *p* = .024. Subjective pain was also associated with the anxiety symptoms, *B* = 2.79, *SE* = 1.08, *t*(65) = 2.57, *p* = .012. These symptoms were significantly associated with the COMM score, *B* = .41, *SE* = .08, *t*(65) = 5.04, *p* < .001. Finally, the relationship between the subjective pain and the COMM score disappeared (*B* = .91, *SE* = .79, *t*(64) = 1.15, *p* = .252), when controlling for anxiety, *B* = .38, *SE* = .09, *t*(64) = 4.47, *p* < .001. However, in men, it is not possible to confirm significant mediation since the necessary prior criteria were not met. Thus, as seen in the previous section, the relationships between pain and opioid consumption (*p* = .087), and between pain and symptoms of anxiety (*p* = .081), were not statistically significant.

#### Depression symptoms mediate between subjective pain and opioid consumption

As indicated in the previous analysis, subjective pain was linearly related to the COMM score. Second, subjective pain was also associated with the depression symptoms, *B* = 1.09, *SE* = .27, *t*(95) = 3.94, *p* < .001. Third, these symptoms were significantly associated with COMM *B* = 1.34, *SE* = .18, *t*(95) = 7.40, *p* < .001. Finally, the relationship between the subjective pain and the COMM score is not significant, *B* =.54, *SE* = .57, *t*(94) = .94, *p* = .347, when controlling for depression, *B* = 1.26, *SE* = .19, *t*(94) = 6.51, *p* < .001. Furthermore, the Sobel test indicated that this mediation was also significant, *z* = 3.44, *p* < .001.

As in the previous section, this mediation model was replicated separately for women and men. In women, very similar results were obtained, indicating that depression mediated the relationship between perceived pain and COMM score (Sobel Test: *z* = 2.8, *p* = .004). So, as previously reported, subjective pain was significantly associated with the COMM score. Subjective pain was also associated with the depression symptoms, *B* = 1.25, *SE* =.38, *t*(65) = 3.23, *p* = .002. These symptoms were significantly associated with the COMM score, *B* = 1.24, *SE* = .21, *t*(65) = 5.73, *p* < .001. Finally, the relationship between the subjective pain and the COMM score disappeared, *B* = .51, *SE* = .79, *t*(64) =.64, *p* = .52, when controlling for depression, *B* = 1.19, *SE* = .23, *t*(64) = 5.06, *p* < .001. In contrast, in men these results are not replicated. it is also not possible to speak of significant mediation for men since the necessary prior criteria were not met. The relationships between pain and COMM score (*p* = .087), and between pain and symptoms of depression (*p* = .074), were not statistically significant.

#### Anhedonia symptoms mediate between subjective pain and opioid consumption

As indicated in the previous analysis, subjective pain was linearly related with the COMM score. Second, subjective pain was also associated with the anhedonia symptoms, *B* = .287, *SE* = .12, *t*(95) = 2.31, *p* = .023. Third, these symptoms were significantly associated with the COMM score, *B* = 2.15, *SE* = .47, *t*(95) = 4.54, *p* < .001. Finally, the relationship between the subjective pain and the COMM score was debilitated, *B* = 1.44, *SE* = .59, *t*(94) = .94, *p* = .016, when controlling for anhedonia, *B* = 1.88, *SE* = .47, *t*(94) = 3.96, *p* < .001. Furthermore, the Sobel test indicated that this mediation was also significant, *z* = 2.11, *p* < .017. In this case, it was not possible to replicate the mediation analyses by separating the data for women and men because the first necessary criterion for mediation was not met (see Figure 4 for all criteria). Specifically, for both women and men, there was no significant linear relationship between subjective pain (IV) and anhedonia (M), which prevents further examination of the other relationships.

**Figure 4.**
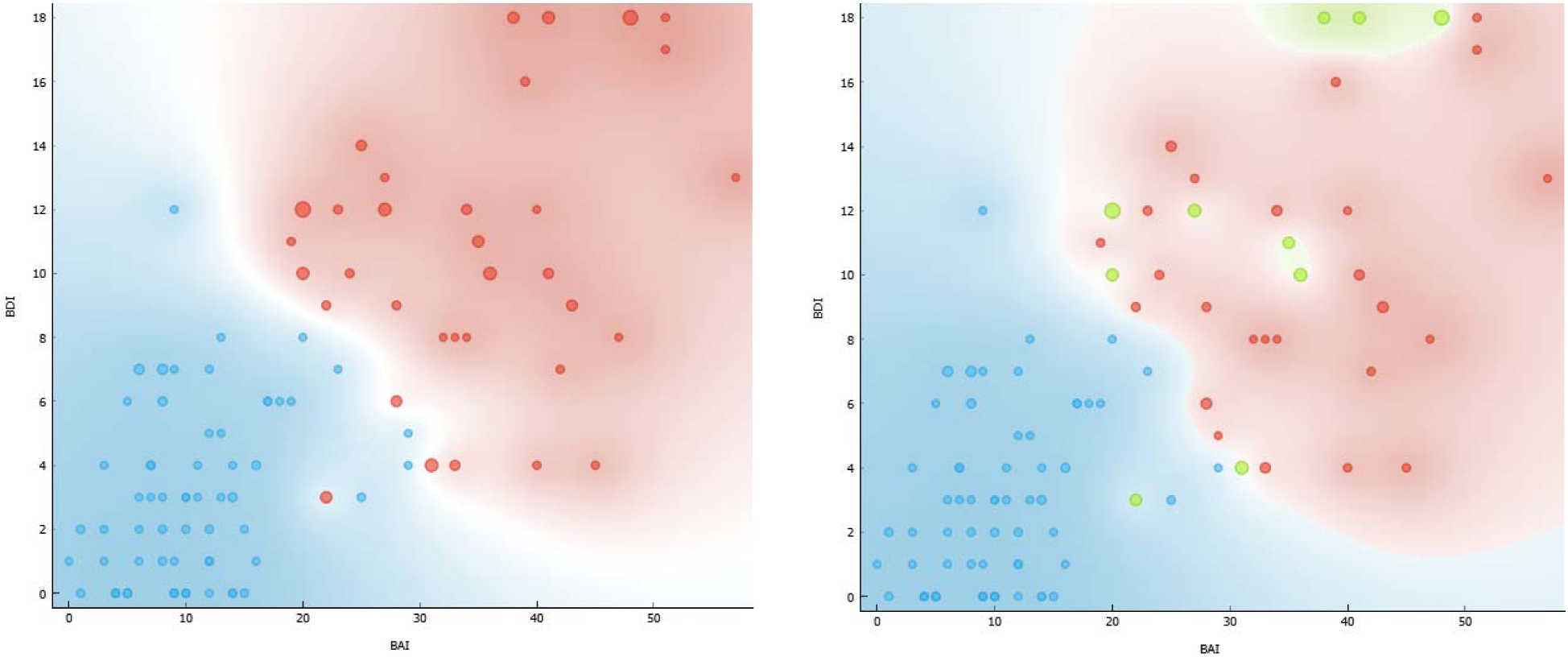
Affective state clusters Anxiety scores (BAI) on the x-axis and depression scores (BDI) on the y-axis. The size of the points represents the level of anhedonia (SHAPS): the larger the size, the higher the anhedonia. On the left, the 2-cluster solution: in red, the highest levels of anxiety, depression, and anhedonia; in blue, the lowest levels. On the right, the 3-cluster solution, with a similar representation but distinguishing (in green) the patients who have high anxiety and depression, but also anhedonia.

### Cluster analysis for affective states and their differences in pain and opioid consumption

Complementarily, the following block of analysis focused on the extraction of clusters (using the k-means method) based on the similarity or distance of patients’ scores in anxiety, depression, and anhedonia. The silhouette score revealed that the optimal solution was between 2 clusters (silhouette score = .504) and 3 clusters (silhouette score = .486). As shown in Figure 4, the 2-cluster solution identified one group with impaired affective states (high anxiety, depression, and anhedonia; negative affect) and another with moderate or mild levels (neutral affect). On the other hand, the 3-cluster solution still showed a similar division but distinguished between those patients who, although both having high anxiety and depression, may or may not have high levels of anhedonia.

Subsequently, the levels of subjective pain and opioid consumption were compared among the extracted clusters. Gender was included in the MANOVAs without reporting a significant effect. Focusing first on the 2-cluster solution, as shown in Table 5, the group with negative affective states scored significantly higher in subjective pain (both over the last month and the last day) and reported particularly high score in the COMM, indicating increased risk of opioid misuse. In contrast, regarding the 3-cluster solution, patients with negative affective states who also reported high anhedonia were found to be midway between the group with negative affect without anhedonia and the group with a more neutral affective state in terms of their subjective pain. This group did not show significant differences from either of the other two groups, although clear differences were observed among the groups. Lastly, risk of opioid misused in the 3-cluster solution was similar to that in the 2-cluster solution, where both groups with negative affective states (with and without anhedonia) reported higher score in COMM compared to the group with a neutral affective state.

**Table 5.** Differences in subjective pain (Last month and day VAS), and opioid consumption (COMM) by affective states (2 and 3 cluster solutions)

| *2 clusters-solution* | Neutral affect (64) | Negative affect (33) | | *F* | df<br>intra | df<br>error | *p*-value | $\eta_p^2$ |
| --- | --- | --- | --- | --- | --- | --- | --- | --- |
| Last-month VAS | 7.38 ± 1.84 | 8.55 ± 0.95 | - | 12.04 | 1 | 95 | .001*** | .062 |
| Day VAS | 6.55 ± 2.07 | 7.72 ± 2.16 | - | 6.79 | 1 | 95 | .011* | .043 |
| COMM | 8.56 ± 6.11 | 22.50 ± 12.16 | - | 57.25 | 1 | 95 | <.001*** | .347 |
| *3 clusters-solution* | Neutral affect (63) | Negative affect<br>without anhedonia (25) | Negative affect<br>with anhedonia (9) | *F* | df<br>intra | df<br>error | *p*-value | $\eta_p^2$ |
| Last-month VAS | 7.37 ± 1.86<sup>a</sup> | 8.62 ± 1.01<sup>b</sup> | 8.35 ± 0.74<sup>a b</sup> | 6.09 | 2 | 94 | .003** | .063 |
| Day VAS | 6.51 ± 2.06<sup>a</sup> | 8.12 ± 1.82<sup>b</sup> | 6.85 ± 2.68<sup>a b</sup> | 5.34 | 2 | 94 | .006** | .051 |
| COMM | 8.40 ± 6.01<sup>a</sup> | 21.88 ± 11.35<sup>b</sup> | 23.70 ± 14.03<sup>b</sup> | 29.63 | 2 | 94 | <.001*** | .364 |

Mean ± SD by affective states clusters (n); df, degrees of freedom; groups with the same letter in superscript (<sup>a</sup> or <sup>b</sup>) do not differ significantly; different letters represent significant differences according to *post hoc* tests; \* significance at the .05 level; \*\* significance at the .01 level; \*\*\* significance at the .001 level

## Discussion

The present study demonstrates that the variables defining affective states (BAI, BDI, and SHAPS) are positively correlated with subjective pain levels (VAS for the last month and VAS for the day) and with opioid misuse (COMM) for the first time in a Spanish clinical sample. Additionally, anxiety and depression levels show a significant correlation within this population of chronic pain patients. When analyzing these correlations by sex, both men and women exhibit clear associations between anxiety-depressive symptomatology and patterns of opioid misuse. However, only women show correlations between subjective pain and anxiety-depressive symptomatology. Furthermore, comparative analyses based on the severity of anxiety-depressive symptomatology indicate that patients with moderate to severe levels of these symptoms presented higher COMM scores, suggesting a higher risk of prescription opioid misuse. Lastly, no significant relationship was found between these variables and alcohol consumption in the studied population.

The present study highlights a clear association between subjective pain levels and anxiety-depressive symptomatology. The link between anxiety and depression and perceived pain in chronic pain patients has been well-documented over the past decades [12,30,56], underscoring the importance of incorporating these variables into the diagnosis and treatment of chronic pain, particularly from a biopsychosocial perspective. One of the most extensively studied symptoms in mood disorders is anhedonia, which, as in prior studies [15,58], was found to be associated with perceived pain levels in the present work. Additionally, our findings suggest that anhedonia is also linked to misuse of prescription opioid medication, indicating a close relationship between this symptom and subjective pain perception. A critical variable to consider is sex, as substantial evidence points to sex differences in pain perception, the analgesic properties of treatments, and the prognosis of pain-related conditions [19,25,50,52]. In this regard, the current study shows that only women exhibit a significant correlation between perceived pain and anxiety-depressive symptomatology. These findings emphasize the importance of considering sex differences when developing diagnostic and treatment strategies for chronic pain, ensuring that interventions are tailored to specific populations.

As previously documented, pain-induced negative affective states are strongly associated with substance use [60,63]. Extensive research has shown that patients with chronic pain who have been diagnosed with OUD or AUD exhibit significantly higher rates of anxiety and depression compared to the general population [4,7,14,29,38,41,64]. Furthermore, the simultaneous use of alcohol and opioids can hinder the management of chronic pain [36]. Surprisingly, 16-25% of patients treated for chronic pain exhibit risky alcohol intake what increase the risk of developing OUD [26,31,32,39,62]. This body of evidence highlights the intricate relationship between chronic pain, negative affect, and psychiatric comorbidities, further emphasizing the importance of addressing these factors in the management of chronic pain patients but also the prevention of opioid misuse in the clinical setting.

The present study did not find a significant relationship between anxiety-depressive or perceive pain symptomatology and alcohol risk consumption as measured by the AUDIT. It is important to consider the specific characteristics of these clinical sample. Firstly, patients under medical supervision in the pain unit are typically advised to avoid alcohol consumption due to its potential interactions with prescribed medications, which could pose significant health risks, including an increased risk of opioid overdose [16]. Additionally, these patients, who report particularly low levels of alcohol consumption, may be hesitant to disclose alcohol use that contravenes medical advice, or they may rely on opioids as a self-treatment mechanism, reducing the need to turn to alcohol as a coping strategy. Alcohol consumption as a means of alleviating negative affective states or as a coping strategy is one of the mechanisms involved in the initiation and maintenance of binge drinking behaviors [22,33]. To further explore the unexpected lack of correlation in this study, alternative approaches to measuring alcohol consumption should be considered. One possible solution could be the combination of the AUDIT with questions designed to calculate alcohol intake in terms of standard drink units, as social acceptance of alcohol may lead patients to underreport their consumption, resulting in artificially low AUDIT scores. Incorporating blood or urine alcohol measurements into future research could also enhance the validity and reliability of self-reported alcohol use.

Our population of chronic pain patients demonstrates a pattern of risk for prescription opioid misuse, with a mean score exceeding 13 on the COMM scale, surpassing the established cut-off of 9 for identifying risk behaviors. Notably, this risk of opioid misuse is positively correlated with higher scores in anxiety-depressive symptomatology, with no significant differences observed between men and women. However, regarding the sex variable, the relationship between negative affective states and opioid misuse in chronic pain patients remains inconclusive, as existing literature presents heterogeneous findings [28,42,54]. It is well-documented that negative affective states modulate the efficacy of opioid treatments, with the presence of anxiety, depression, and anhedonia reducing opioid treatment effectiveness [20]. In this study, we explored whether anxiety-depressive symptomatology mediates the relationship between subjective pain levels and opioid misuse risk. Consistent with prior findings, we found that symptoms of anxiety, depression, and anhedonia mediate this relationship. Interestingly, sex-specific analyses revealed that this mediation effect is present only in women. As previously discussed, sex differences in pathophysiology, treatment efficacy, and comorbidities are well-established [19,25,50,52]. Taken together, the data underscores the clear relationship between pain severity, negative affective states, and risk of misuse of prescription opioids. Therefore, it is imperative to continue investigating these relationships to develop preventive strategies and targeted holistic therapies that address these issues. Psychological therapies have shown promise in alleviating both the physical and psychological symptoms associated with chronic pain, offering the potential to improve the quality of life for these patients [49].

Therefore, the identification of anxiety-depressive symptomatology in chronic pain patients emerges as a valuable tool for detecting and addressing potential opioid use disorders (OUDs) in this population. The presence of these negative affective states is among the most significant risk factors for substance use and relapses in substance use disorders [7,24,34,35,38]. To define a risk profile for these patients, a cluster analysis of negative affective states was conducted. This analysis revealed that patients within clusters characterized by negative affective states, with or without anhedonia, exhibit a higher risk of opioid misuse compared to those without such symptomatology. These findings highlight the critical need for developing targeted interventions for chronic pain patients with anxiety-depressive symptomatology to detect and manage the negative affective states associated with pain and opioid misuse. Additionally, such interventions can play a key role in preventing the development of opioid misuse patterns. Indeed, chronic pain patients who integrate psychological therapies into their standard treatment regimens demonstrate improved quality of life, enhanced pain relief, reduced risk of disability, and decreased fear-avoidance behaviors [49]. Thus, highlights that physical and psychological variables influence pain pathologies and associated comorbidities. Consequently, addressing both the psychological and physical dimensions of chronic pain through comprehensive treatment strategies may significantly mitigate the risks associated with opioid misuse.

This study shows important relationships between anxiety, depression and subjective pain, and opioid misuse, without showing any relationship between alcohol and the previously mentioned variables. However, it is not free of limitations. Among the limitations we must highlight that the number of men in this study is very low, so the differences between sexes may be biased for this reason. In addition, AUDIT scores, measures of drinking patterns, were extremely low, and we have no other measure to assess the validity of these results. For this reason, the alcohol results should be treated with great caution. It is also important to note that this study focused on a population under close supervision by a specialized pain unit. Most individuals with chronic pain are often managed by other specialties or in primary care settings, which may influence the treatment they receive and, consequently, the variables measured in this study. Furthermore, different types of pain have been included in the analysis, which could give rise to different psychological patterns in relation to anxious-depressive symptomatology. Finally, as this is a cross-sectional study, we cannot affirm a causal relationship between variables, but we can establish relationships between them, which can be used in future longitudinal studies to unravel causal relationships between these variables.

In conclusion, this study underscores the critical interplay between anxiety-depressive symptomatology, subjective pain levels, and the risk of opioid misuse in chronic pain patients. The strong correlation between negative affective states and opioid misuse, particularly in women, highlights the importance of integrating psychological assessments into the management of chronic pain. These findings align with previous research, reinforcing the necessity of a biopsychosocial approach to chronic pain treatment. The lack of correlation between anxiety-depressive symptomatology and alcohol misuse may be attributed to the unique clinical oversight in this patient population, suggesting the need for alternative measures of alcohol consumption in future research. Given the clear influence of negative affective states on opioid treatment efficacy, it is essential to further investigate sex-specific differences and develop tailored interventions. Psychological therapies, shown to improve quality of life and reduce both physical and psychological symptoms, should be prioritized as a fundamental component of pain management strategies. This comprehensive approach could substantially reduce opioid misuse risk and enhance overall patient outcomes.

## Data Availability

All data produced in the present study are available upon reasonable request to the authors

## Acknowledgements

This study was funded by Delegación de Gobierno para el Plan Nacional sobre Drogas (refs. PND2019-I038 and PND2024-I035. We would like to thank the Pain Care Unit of the Hospital General Universitario de Valencia.

